# CrosSplice: A Pipeline for Identifying Rare Splice-Site Creating Variants from Cross-Tissue Transcriptome Data

**DOI:** 10.64898/2025.12.21.25342246

**Authors:** Yuki Yano, Ai Okada, Misaki Ono, Raúl N. Mateos, Tojo Nakayama, Yuichi Shiraishi

## Abstract

**Background:** Despite their profound impact on patients’ lives, most rare and intractable diseases still lack established treatments. Genomic variants that disrupt normal splicing by creating novel splice sites (splice-site creating variants, SSCVs) substantially contribute to the pathogenesis of those conditions. Deep intronic SSCVs are particularly amenable to antisense oligonucleotide (ASO)-mediated splice modulation, yet many of them remain undetected by conventional genomic analyses. Existing approaches to identify SSCVs, including splicing quantitative trait loci (sQTL) analyses and machine learning-based methods, each show limitations in sensitivity or accuracy, hindering the comprehensive identification of clinically actionable SSCVs.

**Methods:** We developed CrosSplice, a novel pipeline that integrates machine learning–based predictions with statistical association testing to robustly identify SSCVs. By leveraging cross-tissue transcriptome data and aggregating splicing signals across multiple tissues, CrosSplice enables comprehensive detection of SSCVs, including rare and tissue-specific variants often missed by conventional methods.

**Results:** Applying CrosSplice to the GTEx dataset (8,656 transcriptomes of 54 tissues from 479 postmortem donors), we identified 1,743 significant SSCVs, 65% of which were deep intronic. Among these, 185 SSCVs were listed in ClinVar, including five pathogenic or likely pathogenic variants. CrosSplice also discovered a novel deep intronic SSCV in *PLA2G6*, the gene responsible for infantile neuroaxonal dystrophy. We experimentally confirmed that ASOs successfully corrected the aberrant splicing pattern induced by this variant.

**Conclusion:** CrosSplice substantially extends the detectable landscape of SSCVs by capturing rare and tissue-specific variants, uncovering pathogenic and therapeutically actionable SSCVs that are frequently overlooked by existing methods. The resulting SSCV catalogue provides a platform for systematic discovery of ASO targets and advances opportunities for precision therapies in rare and intractable diseases.

## Background

Rare and intractable diseases affect an estimated 263-446 million people worldwide(1), imposing a significant societal and economic burden. Approximately 70% of these diseases are thought to be genetic disorders(1). Although recent breakthroughs in genome analysis technologies and nucleic acid therapeutics are paving the way for development of treatments for these diseases, particularly in the neurological field(2–5), the majority of them still lack established therapeutic options.

For patients with certain classes of genomic variants, splice-switching antisense oligonucleotides (ASOs) offer a promising therapeutic strategy restoring functional protein levels(3,4). ASOs are a key modality in nucleic acid therapeutics noted for high specificity and precision in binding to target gene sequences. These features make them particularly suitable for treatment of ultra-rare diseases characterized by extremely low prevalence and marked genetic heterogeneity(4,5). Among genomic variants, splice-site creating variants (SSCVs), in which variants induce abnormal splicing by generating novel splice-sites, are considered particularly amenable to ASO-mediated splice modulation(5). A representative example is sepofarsen(6), an ASO targeting a SSCV located in a deep intronic region of *CEP290* (c.2991+1655A>G; rs281865192), which causes the inclusion of a cryptic exon and leads to Leber congenital amaurosis 10 [MIM 611755]. Sepofarsen has demonstrated molecular correction of splicing and clinical improvement in patients(6,7), progressing to phase1/2 and 2/3 clinical studies(8,9). Additional examples of ASO therapies targeting SSCVs include those developed for xeroderma pigmentosum(10) [MIM 278760], β-propeller protein-associated neurodegeneration(11) [MIM 300894] and Stargardt disease 1(12) [MIM 248200]. Based on these perspectives, compiling a catalogue of pathogenic SSCVs that may serve as therapeutic targets is expected to facilitate the preemptive development of ASO therapies and help avoid missing the narrow therapeutic window of rare and intractable diseases, given their lethality and rapid progression.

Approaches to detect SSCVs can be broadly divided into several major categories. The first identifies splicing quantitative trait loci (sQTL) from paired genome and RNA-seq data by evaluating statistical associations between genomic variants and splicing changes(13,14). However, these methods exhibit inherently low sensitivity for variants with low allele frequencies. The second group comprises machine learning-based algorithms that predict splicing changes from genomic sequences, such as SpliceAI(15) (2019) and MMSplice(16) (2019). More recently, additional models focusing on tissue-specific splicing have been introduced, including Pangolin(17) (2022), ABSplice(18) (2023) and SpliceTransformer(19) (2024). Although these models have improved prediction accuracy through advances in deep learning techniques, there is still room for improvement in both sensitivity and precision.

Here, we developed a novel pipeline, CrosSplice, for the comprehensive detection of SSCVs using cross-tissue transcriptome data. This pipeline integrates the strengths of both sQTL and machine learning–based approaches, thereby enabling robust identification of rare and tissue-specific splicing events. Furthermore, given that one- to two-thirds of alternative splicing events are tissue-specific(20,21), CrosSplice integrates multi-tissue data for comprehensive SSCV detection. After evaluating CrosSplice’s performance, we applied this pipeline to the GTEx v7 dataset and created a catalogue containing 1,743 SSCVs, some of which we have demonstrated to be pathogenic and may serve as potential targets for splice-switching ASO therapies.

## Methods

### ● Transcriptome data collection and preprocessing

We collected and preprocessed mutli-tissue transcriptome data from the Genotype-Tissue-Expression (GTEx) v7 dataset, as previously described(22). Briefly, we obtained RNA-seq data from 8,656 samples across 54 tissues collected from 479 donors. Those RNA-seq were aligned to the GRCh38-based reference genome obtained from the Genomic Data Commons using STAR (v2.7.9b). After alignment, BAM files were sorted, converted to CRAM format, and indexed using SAMtools version 1.9. In this process, we obtained splicing junction files (more specifically, the SJ.out.tab files produced by STAR).

Genomic variant data, including single nucleotide variants (SNVs) and indels, corresponding to the GTEx v7 transcriptome samples were obtained from the whole genome sequencing (WGS) dataset of GTEx V7 in VCF format. Coordinates of the variants were converted from GRCh37 to GRCh38 using liftover. Then, the resulting VCF file was compressed with bgzip and indexed with tabix (-vcf). Next, all variants were annotated with Ensembl Variant Effect Predictor (VEP), which provided associated information including SpliceAI score and gnomAD allele frequency (v3.1.2). Finally, we filtered down to only SNVs with SpliceAI delta scores (DS_AG or DS_DG, representing the probability of the variant creating a new acceptor or donor site, ranging from 0 to 1)(15) greater than 0.1, and with gnomAD allele frequency (AF) less than 0.01.

### ● CrosSplice pipeline

CrosSplice performs a cross-tissue analysis to evaluate whether splicing alterations predicted by SpliceAI occur at a significantly higher frequency in individuals carrying an SSCV candidate compared to those without the variant. Adopting the notation from a previous study(22), the procedure is as follows.

For each SSCV candidate, we first obtained the predicted position of the primary novel splice-site (SS) from SpliceAI, using the DP_AG or DP_DG field (acceptor or donor gain positions relative to the variant). For this primary novel SS, we then identified the matching splice-site (SS) from the annotated transcript, selecting the nearest upstream donor site in the case of an acceptor gain, or the nearest downstream acceptor site in the case of a donor gain. The primary novel splicing junction (SJ) was defined as the region between the primary novel SS and the matching SS. We also identified the hijacked splicing junction (SJ) and hijacked splice-site (SS). The hijacked SS originally paired with the matching SS but was displaced by the primary novel SS, corresponding to the nearest downstream acceptor (for an acceptor gain) or upstream donor (for a donor gain) relative to the matching SS. The hijacked SJ was defined as the region between the hijacked SS and matching SS (Additional file 1: Figure S1). For the process of selecting a transcript, we first fetched all candidate transcripts, which contained primary novel SS within their region, from the GENCODE annotation file (Release 39). After defining the matching SS, hijacked SS, primary SJ and hijacked SJ for each candidate, we prioritized the candidate transcripts in the following order: MANE Select(23), MANE Plus Clinical and other transcripts, and finally selected the one with the shortest hijacked SJ. MANE flag information was obtained from the NCBI FTP site (https://ftp.ncbi.nlm.nih.gov/refseq/MANE/MANE_human/release_1.0/MANE.GRCh38.v1.0.ensembl_genomic.gff.gz). Splicing consequences were predicted using our in-house pipeline (https://github.com/ncc-gap/juncmut/tree/master). Secondary novel splice-site (SS), which represents an additional splice-site forming a cryptic exon together with the primary SS, were also identified within the same in-house pipeline.

(e.g.) For example, in the case of chr22:16808099 C>T, the predicted SpliceAI DS_AG and DS_DG were 0.14 and 0.00, and the corresponding DP_AG and DP_DG were -13 and -16, respectively. Based on these predictions, this variant was classified as “acceptor gain”, with primary novel SS defined as 13nt upstream of the variant (chr22:16808087). Referring to the MANE-select and shortest sized transcript, matching SS was set as the nearest upstream donor site (chr22:16808084) and the hijacked SS as chr22:16825290. Then, hijacked SJ and primary novel SJ were determined as chr22:16808084-16825290 and chr22:16808087-16825290, respectively.

Subsequently, SJ.out.tab files were parsed to count the number of supporting reads for hijacked SJ (#hijacked_SJ) and primary novel SJ (#primary_novel_SJ) for each sample and tissue, and calculated the ratio for a variant as follows:

depth = #hijacked_SJ + #primary_novel_SJ

alternative ratio = #primary_novel_SJ / (depth + 1)

Next, for each tissue, we calculated p-values to measure the difference in the alternative ratio between samples with and without the variant. This calculation was performed using a one-sided Wilcoxon rank-sum test via the wilcoxon.test function in R language. Lastly, we integrated the p-values of each tissue into a single combined p-value using Fisher’s method, which aggregates multiple independent p-values to assess the overall significance across tissues.

### ● sQTL data from GTEx v8

For comparison with SSCVs identified by CrosSplice, we used sQTL data derived from the GTEx v8 dataset and identified using LeafCutter (available at https://storage.googleapis.com/adult-gtex/bulk-qtl/v8/single-tissue-cis-qtl/GTEx_Analysis_v8_sQTL.tar).

### ● Evaluation of false discovery rate by permutation test

To evaluate the empirical false discovery rate (FDR), we performed permutation tests with shuffled pairs of genomic and transcriptome data. The FDR was defined as the proportion of SSCVs expected to arise by chance under permutation. Transcriptome data was randomly reassigned to WGS data containing variant information, thereby disrupting the original genotype–expression correspondence. CrosSplice was applied to these shuffled data pairs, and the number of significant SSCVs detected under the threshold was recorded. Specifically, we defined D^(target)^ as the number of SSCVs identified from the correct combination of datasets and D_i_^(perm)^ as the number of SSCVs obtained from i-th permutation procedure, and calculated the FDR as follows:

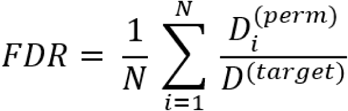

We performed 100 permutation trials in this paper.

### ● Annotation of SSCVs with information on neurodevelopmental disorders

We performed variant annotation using ClinVar (August 2025 release available at https://ftp.ncbi.nlm.nih.gov/pub/clinvar/vcf_GRCh38/clinvar_20250831.vcf.gz), including information on clinical significance, molecular consequence and review status. Also, for genes associated with neurodevelopmental disorders that have the potential to be treated with RNA-based therapeutics, we used a gene list curated by Dutch Center for RNA Therapeutics (DCRT), a nonprofit consortium dedicated to the development of RNA therapies for rare genetic disorders (available at https://irp.cdn-website.com/335a66d0/files/uploaded/DCRT_Gene_list_August_2024.xlsx).

### ● Minigene assay

We selected two variants for validation in vitro, *ABCA7* chr19:1061893 G>C and *PLA2G6* chr22:38115172 C>T, based on low p-values and their association with neurodevelopmental diseases. *ABCA7* (intron 40, exon 41, intron 41, exon 42, intron 42) and *PLA2G6* (intron 13, exon 14, intron 14, exon 15, intron 15) mini-gene constructs were generated in pSE vectors. The constructs were transfected into E. coli DH5α Competent Cells (TOYOBO) and E. coli HST08 Premium Competent Cells using In-Fusion Cloning kit (Takara) for amplification. Minigenes were extracted and purified using HiYield Plasmid Mini Kit (Real Genomic) and NucleoBond Xtra Midi/Maxi Kit (Takara). Before transfection, HEK293T cells were cultured for 24 hours, in a 24 well plate for *ABCA7* and in a 96 well plate for *PLA2G6*. HEK293T cells were then transfected with minigenes using Lipofectamine 3000, and incubated for 48 hours at 37℃. From *PLA2G6* minigene-transfected cells, RNA was extracted, and cDNA was synthesized using Cells-to-CT kit. From *ABCA7* minigene-transfected cells, RNA was extracted with FastGene RNA basic Kit (Nippon Genetics), and cDNA was synthesized using RT Master Mix (Takara). We performed PCR to amplify the target region from cDNA, and evaluated changes in splicing pattern with electrophoresis assay.

### ● Antisense oligonucleotide design and in vitro treatment

Antisense oligonucleotides (ASOs) with full phosphorothioate (PS) + 2’-O-Methoxyethyl (2’MOE) modifications were designed targeting splicing motifs and the variant. Exonic splicing enhancer (ESE) motifs were predicted using ESEfinder(24). Control ASO was also designed to be non-targeting in the human genome. HEK293T cells were co-transfected with 0.3μM ASO and minigene constructs with Lipofectamine 3000, and incubated for 48 hours. RNA extraction, cDNA synthesis, PCR and electrophoresis assay was performed as described above. ASO sequences are listed in Table 1.

**Table 1.**
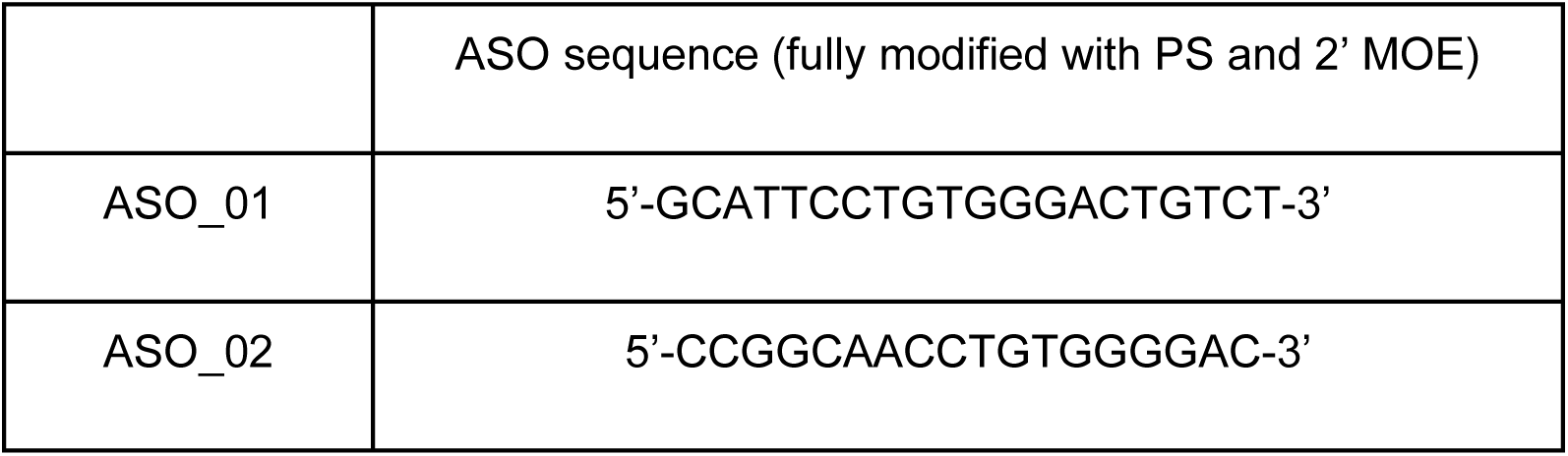
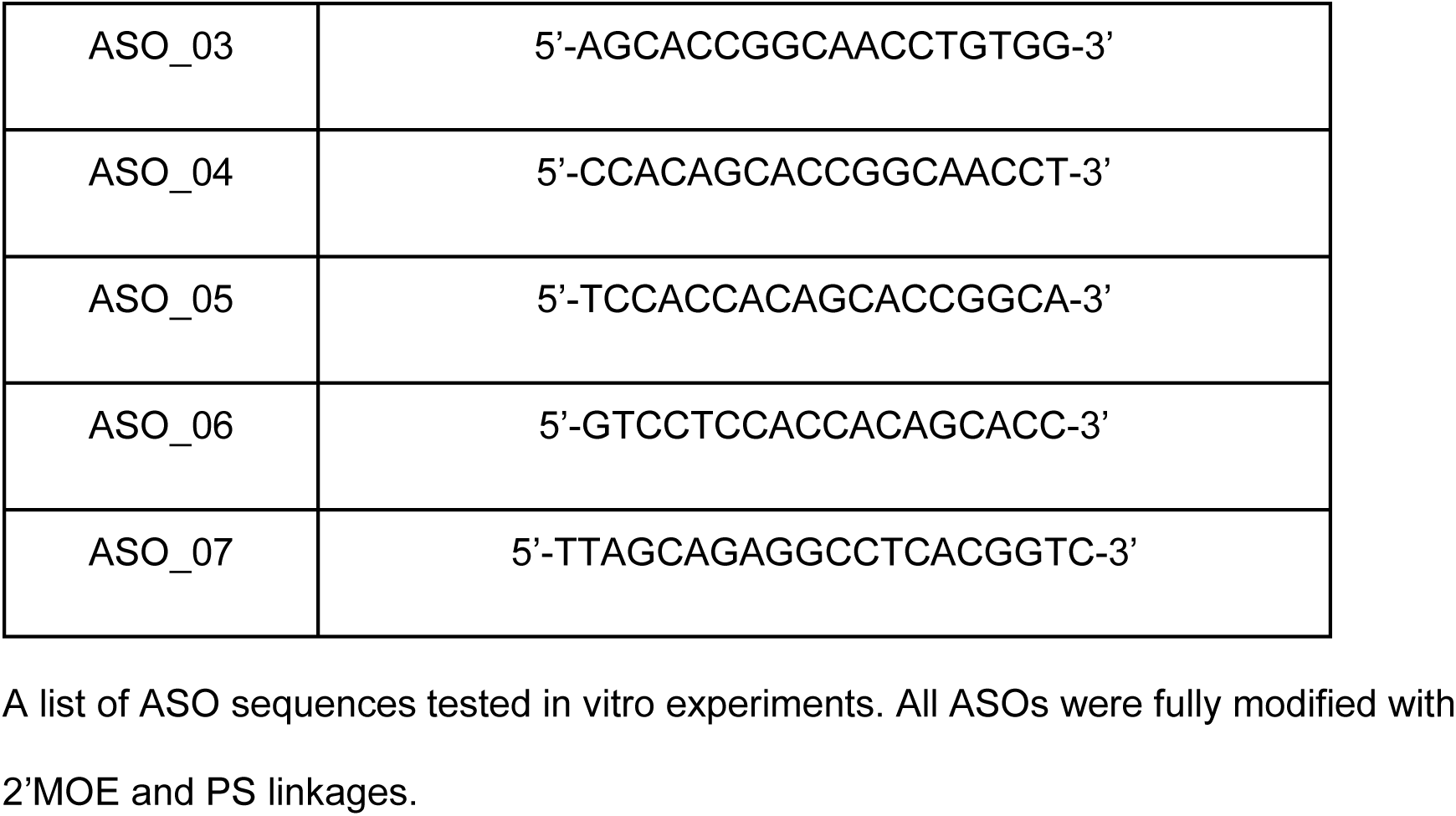
List of ASO sequences.

### ● Data analyses

All analyses were performed in Python 3.12.0 and R 4.3.3. Figures were generated using the ggplot2 and UpSetR package in R, and the matplotlib package in Python.

### ● Use of generative AI and AI-assisted technologies in the writing process

During the preparation of this work the authors used ChatGPT-5 (OpenAI) in order to improve the clarity and readability of the manuscript text. After using this tool, the authors reviewed and edited the content as needed and take full responsibility for the content of the publication.

## Results

### ● Method overview

CrosSplice initially filters candidate variants by AF and SpliceAI delta score. It focuses on rare variants with AF ≤ 0.01, as common variants are generally considered less likely to be pathogenic, particularly in the context of rare diseases(25). In addition, CrosSplice considers SNVs with even minimal SpliceAI delta scores for acceptor gain or donor gain (DS_AG or DS_DG ≥ 0.1). This threshold is more permissive than the “high recall” threshold (0.2) noted in the SpliceAI documentation (SpliceAI GitHub. https://github.com/Illumina/SpliceAI), and was selected to capture a broader range of potential SSCVs. Here, for each candidate SSCV, we used DP_AG and DP_DG predicted by SpliceAI to infer a newly created splice-site (primary novel SS) and resulting primary novel SJ, spanning the primary novel SS and its corresponding matching SS. In addition, for each primary novel SJ, we identified the corresponding hijacked SJ that would normally be utilized but was displaced by the SSCV (Figure 1a).

**Figure 1.**
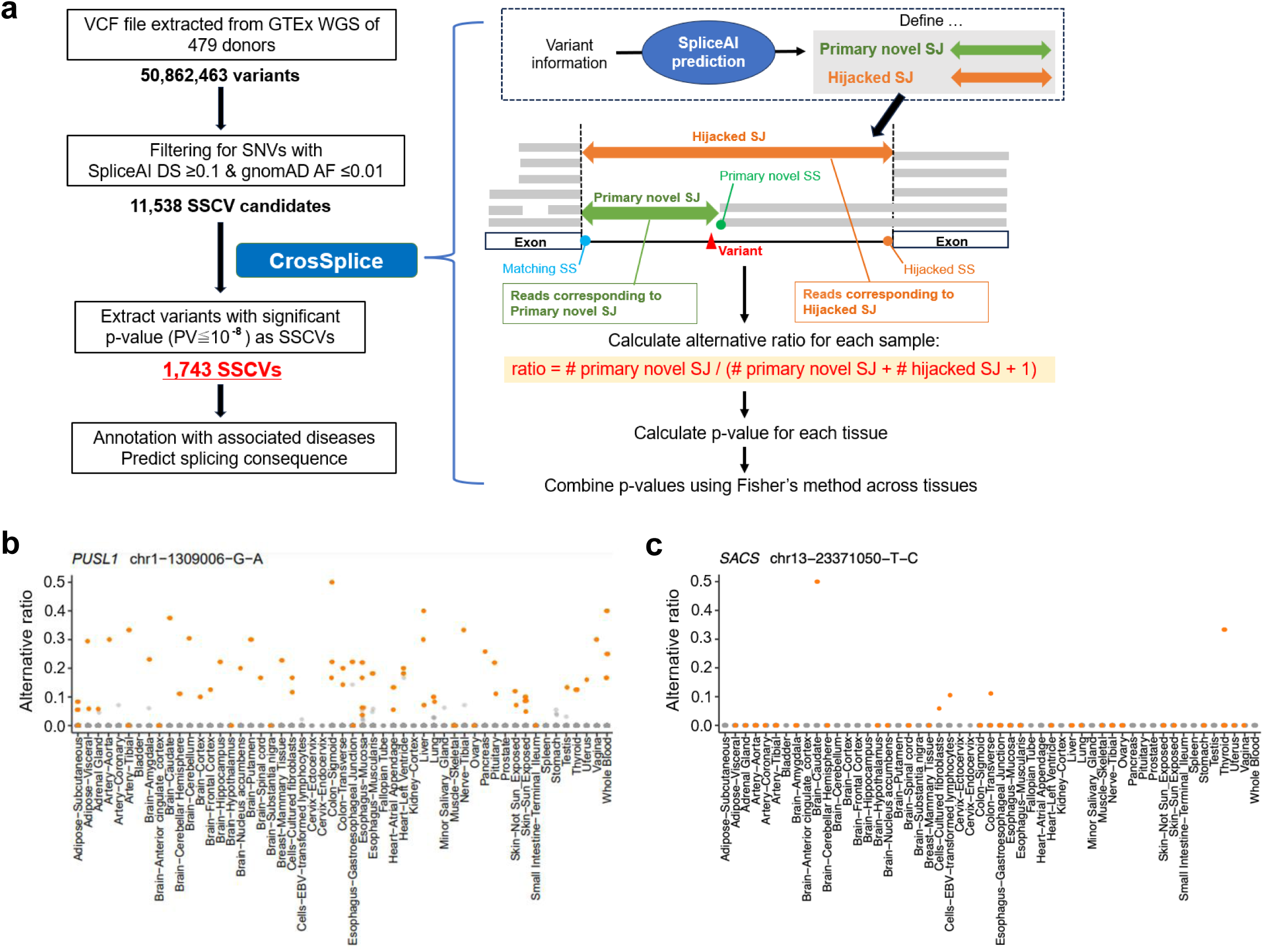
Overview of CrosSplice pipeline and representative examples of SSCVs with tissue-wide splicing alterations. (a) Workflow of CrosSplice pipeline. (b) (c) Alternative ratios across tissues for (c) chr1:1309006 G>A and (c) chr13:23371050 T>C. Orange dots indicate alternative ratios in samples harbouring the variant, and grey dots represent those in samples without the variant. A greater separation between two groups indicates a stronger likelihood that the variant genuinely causes the predicted splicing change.

Next, CrosSplice evaluates whether each SSCV induces predicted abnormal splicing using transcriptome data from the GTEx project, which encompasses a wide range of human tissues. For this purpose, we calculated “alternative ratio”, defined as #primary_novel_SJ / (#primary_novel_SJ + #hijacked_SJ + 1), and examine whether samples carrying an SSCV tend to show higher alternative ratios across tissues (Figure 1b,c). For each tissue, the significance of differences in the alternative ratio between samples with and without the SSCV was assessed using the Wilcoxon rank-sum test. The resulting p-values from all available tissues are then aggregated using Fisher’s method, representing the overall effect of each SSCV. The combined p-values are converted to ‘CrosSplice score’ (–log10 combined p-value), which quantifies the splicing impact of SSCVs across tissues.

The novelty of CrosSplice lies in its ability to integrate machine learning–based predictions with tissue-wide statistical evaluation. Unlike conventional sQTL approaches that unbiasedly test a vast number of variant-splicing associations, CrosSplice restricts its analysis to variants and their consequences predicted by SpliceAI with high prior probabilities. By narrowing the hypothesis space to events prioritized by machine learning, this strategy markedly improves the accuracy of SSCV detection. Moreover, by aggregating splicing signals across multiple tissues, CrosSplice enables tissue-independent yet comprehensive detection, increasing the power to identify rare variants with consistent splicing effects across tissues.

### ● Evaluation of CrosSplice with GTEx project dataset

We evaluated the CrosSplice pipeline using GTEx WGS data of 479 donors (Figure 1a). From 50,862,463 variants, annotation and filtering based on SpliceAI delta score and gnomAD AF yielded 11,538 SSCV candidates. We performed CrosSplice on these candidates and identified 1,743 SSCVs with CrosSplice scores ≥ 8 (the complete list of significant SSCVs is provided in Additional file 1: Table S1). Among the 1,743 significant SSCVs, 758 variants (43.5%) were those creating acceptor sites and 985 variants (56.5%) were creating donor sites. We also evaluated CrosSplice in a single-tissue setting using clinically accessible tissues: *Whole Blood* and *Skin-Sun Exposed* (tissue names are shown according to the original GTEx nomenclature). Compared with the multi-tissue analysis, the single-tissue analyses identified fewer SSCVs: 290 in *Whole Blood* and 338 in *Skin-Sun Exposed* (Figure 2a). Nevertheless, most of the single-tissue SSCVs overlapped with those identified in the multi-tissue analysis, accounting for 93.4% of SSCVs from *Whole Blood* and 95.9% from *Skin-Sun Exposed*. The two-tissue analysis (*Whole Blood* and *Skin-Sun Exposed*) yielded 349 SSCVs and showed an overlap of 95.4%.

**Figure 2.**
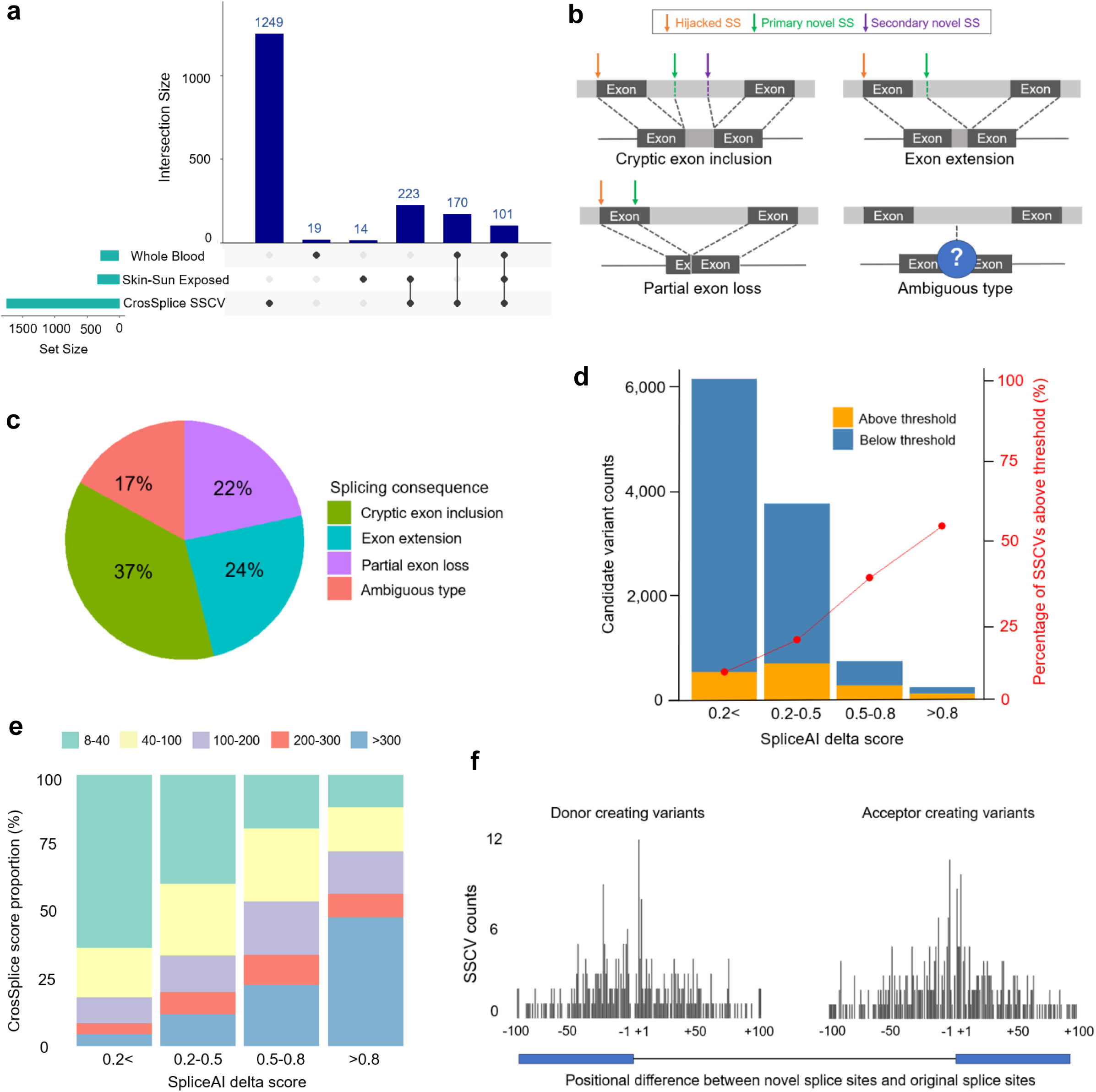
Statistical properties and splicing consequence patterns of SSCVs detected by CrosSplice. (a) UpSet plot showing the overlaps between SSCVs identified from individual single tissues (*Skin-Sun Exposed* and *Whole Blood*) and SSCVs detected in the multi-tissue analysis by CrosSplice. Note that the intersection between *Skin–Sun Exposed* and *Whole Blood* was not separately shown, as it completely overlapped with the intersection including the SSCVs identified in the multi-tissue analysis. (b) Schematic illustration of four types of splicing consequences. Orange arrows indicate hijacked splice-sites, and green arrows denote primary novel splice-sites generated by SSCVs. In the cryptic exon inclusion pattern, a secondary novel splice-site, indicated by a purple arrow, is additionally generated. Ambiguous type contains unclassifiable cases due to the variants’ intergenic locations or insufficient read support to determine splicing patterns. (c) Proportions of splicing consequence types among 1,743 SSCVs identified by CrosSplice. (d) Counts of candidate SNVs and SSCVs stratified by SpliceAI delta score ranges. Orange segments indicate SSCVs, defined as variants exceeding the CrosSplice significance threshold (≥ 8). Red dots represent the proportion of SSCVs within each delta score range. (e) Distribution of CrosSplice scores in each SpliceAI delta score range. Bars are colored according to the corresponding CrosSplice score, as shown in the legend. (f) Distribution of the relative positions of primary novel splice-sites with respect to the hijacked splice-sites. The x-axis indicates the nucleotide distance from the exon-intron boundary.

We further classified SSCVs into four categories based on their splicing consequences: 644 (37%) were cryptic exon inclusion, generating a novel exon within an intron; 378 (22%) were partial exon loss, truncating an existing exon; 425 (24%) were exon extension, extending an existing exon; and the remaining SSCVs were classified as ambiguous type (unclassifiable cases due to intergenic locations or insufficient read support; Figure 2b, c).

When comparing SSCVs detected by CrosSplice with sQTLs from the GTEx dataset identified by the conventional sQTL approach (see Methods for details), we found only seven overlapping variants (Additional file 1: Table S2). This limited overlap likely reflects a fundamental difference in detection scope: CrosSplice specifically targets rare variants (AF ≤ 0.01) with strong splicing effects, whereas conventional sQTL analyses mainly capture common variants with modest regulatory effects(26–28) (Additional file 1: Figure S2). We also estimated FDR by performing 100 permutation tests with shuffled combinations of WGS and RNA-seq data. Under the significance threshold used in this study (CrosSplice score ≥ 8), the FDR was 5.2%. More stringent thresholds yielded even lower FDRs (≥ 10: 4.3%, ≥ 12: 3.6%; Additional file 1: Table S3).

### ● Relationship between SpliceAI and CrosSplice scores, and positional characteristics of SSCVs

We investigated the relationship between CrosSplice scores and SpliceAI delta scores. Variants with higher SpliceAI delta scores more frequently exceeded the CrosSplice significance threshold (Figure 2d), and CrosSplice scores also increased with higher SpliceAI delta scores (Figure 2e). Although the proportion of variants surpassing the threshold was low in the low- and mid-range of delta scores (< 0.2 and 0.2–0.5), a substantial number of SSCVs were still detected due to the large number of candidate variants in those score ranges. Specifically, 620 SSCVs in the < 0.2 range remained after applying CrosSplice, and 26 of them exhibited very high CrosSplice scores (> 300), including 20 variants located in deep intronic regions (defined as positions more than 100 bp away from exon-intron boundaries; Additional file 1: Figure S3).

Next, we evaluated the distribution of novel splice-sites relative to exon-intron boundaries identified by CrosSplice. Approximately 65% of SSCVs were located within deep intronic regions, while the remaining variants were predominantly enriched near exon-intron boundaries (Figure 2f).

### ● Intron length influences splicing consequences via intron/exon definition modes

When focusing on the length of introns affected by SSCVs (corresponding to hijacked SJs in CrosSplice), we observed clear tendencies in splicing consequences (Figure 3a). Partial exon loss was most prevalent in the shortest introns (1-250 bp), whereas exon extension was dominant in the intermediate length range (250-1200 bp). In even longer introns, cryptic exon and ambiguous pattern were the most frequently observed (Figure 3b-d). Among the SSCVs causing partial exon loss in shortest introns (1-250bp), 85% were exonic variants that create novel splice-sites, whereas the remaining 15% were primarily intronic variants near the boundaries that directly disrupt original splicing motifs (e.g. Figure 3e). These trends can partly be explained by exon- and intron-definition modes of splice-site recognition (Additional file 1: Figure S4). Previous studies have proposed that splice-sites flanked by introns longer than ∼250 bp are recognized via exon definition, whereas those flanked by shorter introns are recognized via intron definition(29,30). These definition modes also influence splice-site selection: when multiple candidate splice-sites exist, the ones located closer to the center of the intron are preferentially utilized (so called “intron-centric proximity rule”), and this preference is especially pronounced in splice-sites with intron definition(29,31). Subsequently, the canonical splice-site is generally located closest to the center of the intron, consequently leaving alternative splice-sites within the exon. Thus, when an SSCV weakens the strength of a canonical splice-site, as illustrated in Figure 3e and S4, an alternative splice-site located within the exon is often utilized, resulting in partial exon loss. This hypothesis is consistent with our results, which showed that the partial exon loss was most frequently observed in shortest introns, where spice sites are preferentially recognized by intron definition.

**Figure 3.**
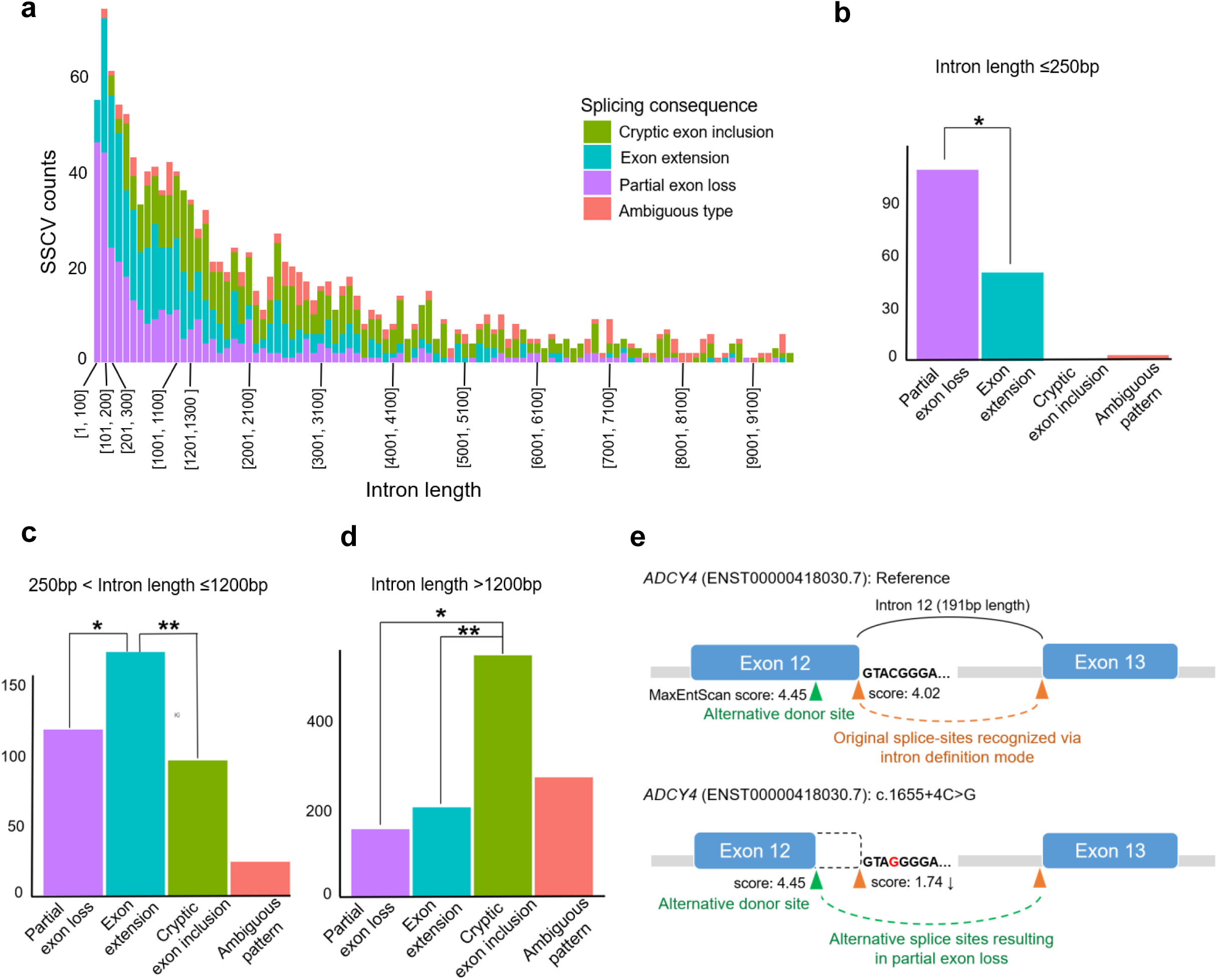
Intron length–dependent patterns of splicing consequences induced by SSCVs. (a) Bar graph showing the distribution of splicing consequence types across intron length categories. This figure displays data for introns with lengths up to 10,000 bp. (b) (c) (d) Proportions of splicing consequence types in introns of short (< 250 bp), medium (250-1200 bp) and long (> 1200 bp) lengths. Asterisk denotes statistically significant differences between categories. (b) * = P < 10^-9^ (c) * = P < 10^-4^, ** = P < 10^-7^ (d) * = P < 10^-15^, ** = P < 10^-15^. (e) An example of splicing change pattern in *ADCY4* (ENST00000418030.7):c.1655+4 C>G. Strength of splice-sites was predicted using MaxEntScan(66). Under the normal condition, the canonical donor site is selected due to its proximity to the center of the intron, despite its low MaxEntScan score. When an SSCV weakens this canonical splice-site, an alternative splice-site (located within exon) is utilized instead, resulting in partial exon loss.

### ● SSCVs affecting the onset of human genetic diseases

To identify SSCVs associated with human genetic diseases, we examined their registration status in ClinVar. Of the 1,743 SSCVs identified, 185 were registered in ClinVar, including 5 annotated as pathogenic or likely pathogenic (one of which was confirmed by the expert panel review), 24 as conflicting classifications of pathogenicity, 45 as uncertain significance, and 111 as benign or likely benign. Among these, only 8 SSCVs were explicitly classified as variants related to splicing (“splice acceptor variant” or “splice donor variant”), including 4 pathogenic SSCVs. In contrast, many SSCVs were annotated as “missense variant” (n=68), “intron variant” (n=54), or “synonymous variant” (n=40), indicating that splicing effects of those SSCVs may have been overlooked in previous evaluations. CrosSplice scores were generally higher in pathogenic/likely pathogenic variants and lower in benign/likely benign variants, but the differences did not reach statistical significance (Additional file 1: Figure S5, S6).

Furthermore, we applied a publicly available list of genes curated by the Dutch Center for RNA Therapeutics (DCRT). The list contains 4,844 genes associated with neurodevelopmental disorders that are considered potential targets for RNA-based therapeutics. Based on the list, approximately one-third of SSCVs (a total of 584 SSCVs) affected genes related to neurological disorders. Specifically, 15 SSCVs caused aberrant splicing in nine major genes associated with widely recognized neurological disorders: three SSCVs in *ITPR1* (associated with spinocerebellar ataxia (SCA) 15/16 [MIM 606658](32,33) and SCA29 [MIM 117360](34)), two each in *PLA2G6* (infantile neuroaxonal dystrophy (INAD) [MIM 256600](35) and PARK14-linked dystonia-parkinsonism [MIM 612953](36)), *HTT* (Huntington’s disease [MIM 143100](37)), *ATXN1* (SCA1 [MIM 164400](38)) and *APP* (familial Alzheimer’s disease [MIM 104300](39)), and one each in *SYNJ1* (developmental and epileptic encephalopathy 53 [MIM 617389](40) and PARK20 [MIM 615530](41)), *PARK7* (autosomal recessive early-onset Parkinson disease [MIM 606324](42)), and *NOP56*(SCA36 [MIM 614153](43)).

### ● Validation of splicing effects of SSCVs in vitro experiments

Among the SSCVs detected in the current study, we focused on ENST00000263094.11:c.5570+5 G>C (rs200538373), a variant that causes a 14-bp exon extension in *ABCA7* gene (Figure 4a). *ABCA7* is broadly expressed across tissues including the brain(44), and is implicated in lipid metabolism(45,46) and amyloid-beta homeostasis(47). Both common and rare variants in *ABCA7* are associated with Alzheimer’s disease [MIM 104300], primarily through loss-of-function mechanisms. The variant c.5570+5 G>C has been identified as a risk locus in multiple GWAS studies(44). Although ClinVar currently classifies it as a variant of uncertain significance, several studies have shown that it induces exon extension, leading to the formation of a premature termination codon (PTC) and subsequent degradation via nonsense-mediated decay (NMD)(47,48). Other studies, however, have reported an inconsistent reduction in ABCA7 protein levels in carriers, suggesting that c.5570+5 G>C may require a second genetic hit to exert its deleterious effect(47,49). In the CrosSplice analysis, this SSCV and its resultant splicing change were detected across multiple tissues, with a predominant occurrence in brain tissue (Figure 4b). We replicated a previously reported minigene assay(49) to validate the splicing impact of this SSCV in vitro, and confirmed the 14-bp exon extension in samples harbouring c.5570+5 G>C (Figure 4c).

**Figure 4.**
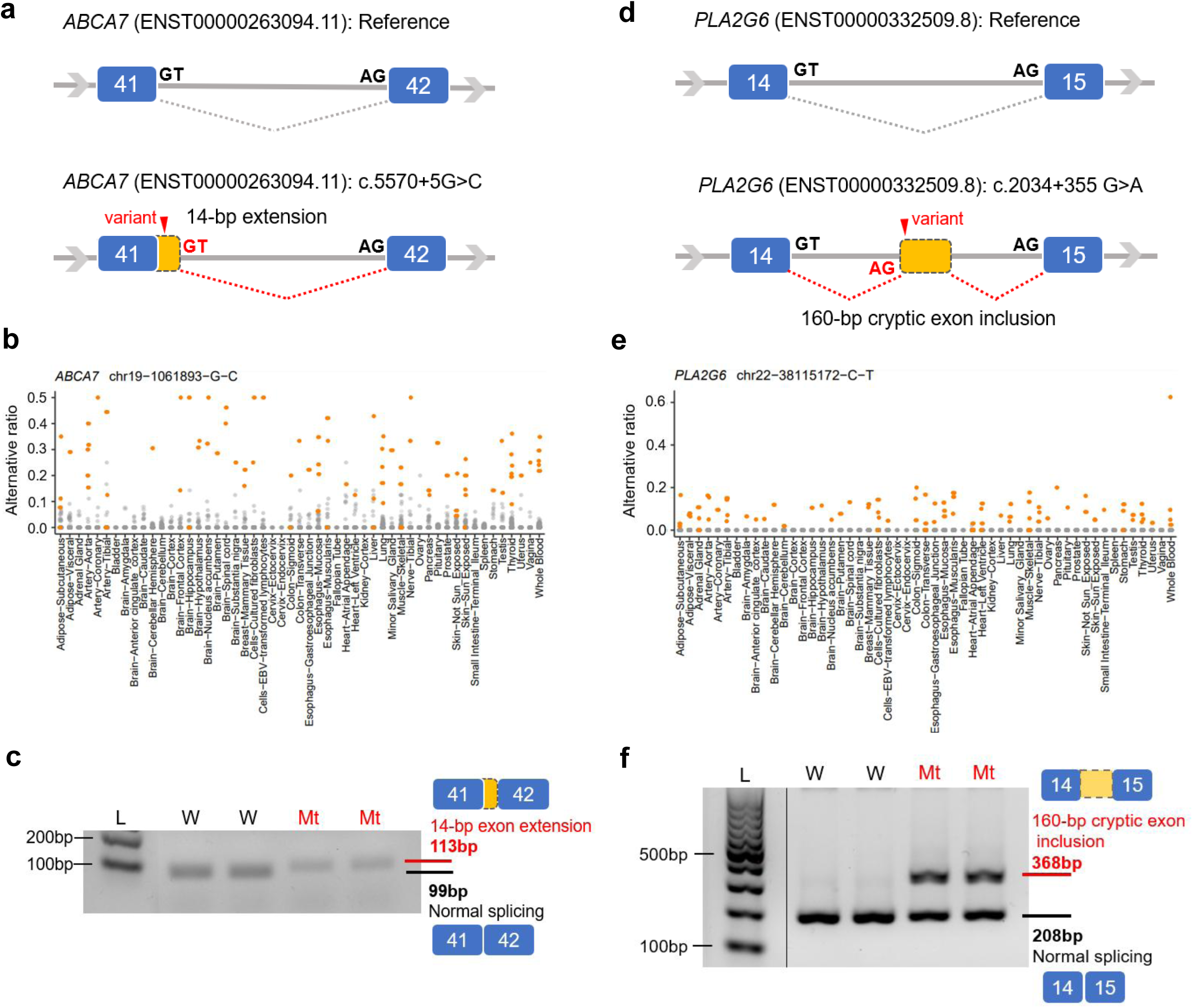
CrosSplice-predicted splicing alterations induced by representative SSCVs in *ABCA7* and *PLA2G6*, validated by minigene assays. (a-c) Splicing alteration induced by *ABCA7* (ENST000263094.11):c.5570+5 G>C. (a) Schematic diagram showing the exon-intron structure and the splicing change. Blue boxes: exons; red arrowheads: SSCVs. (b) Alternative splicing ratios across tissues calculated by CrosSplice. (c) Minigene assay. Mutant type samples (Mt) show a 113-bp band corresponding to the 14-bp exon extension. (d-f) Splicing alteration induced by *PLA2G6* (ENST00000332509.8):c.2034+355 G>A. (d) Exon-intron structure and the splicing change. A newly activated AG motif serves as a novel acceptor site, which pairs with the canonical donor site to form an alternative splice junction. A cryptic splice-site with a GT motif downstream pairs with the canonical acceptor site, resulting in the inclusion of a 160-bp cryptic exon. This cryptic exon inclusion leads to a frameshift, formation of PTC and leading to degradation by NMD. (e) Alternative splicing ratios across tissues calculated by CrosSplice. (f) Minigene assay. Mt samples show a 368-bp band corresponding to the 160-bp cryptic exon inclusion. A lane between the ladder and wild lanes was spliced from the same gel image and rearranged for clarity. A black vertical line indicates where the lane was juxtaposed. L: ladder, W: wild type, Mt: mutant type.

CrosSplice also identified a novel SSCV located in a deep intron of *PLA2G6* gene (Figure 4d, e). The variant, ENST00000332509.8:c.2034+355 G>A (rs147795054), is neither registered in ClinVar nor previously reported as pathogenic. Therefore, its splicing effect and functional impact remained unknown. CrosSplice predicted that the SSCV causes inclusion of a 160-bp cryptic exon, which was verified using a minigene assay (Figure 4f). Inclusion of the 160-bp cryptic exon leads to a frameshift, generating a PTC and subsequent degradation of the transcript via NMD. *The PLA2G6* gene encodes the iPLA2 protein, which is abundant in mitochondrial inner membranes and plays an essential role in maintaining mitochondrial cell membrane homeostasis(35,50). Mutations in *PLA2G6* are known to cause PLA2G6-associated neurodegeneration disorders (PLAN), which includes INAD and PARK14-linked dystonia-parkinsonism, both of which are autosomal recessive and characterized by iron accumulation in the brain(50–53). INAD patients develop normally until the disease onset in infancy, after which they exhibit motor/cognitive regression, pyramidal signs, visual disturbance and cerebellar ataxia(50,52), ultimately leading to respiratory and feeding difficulties. The disease course is typically fatal, with a lifespan of only 5-10 years. Previous studies reporting *PLA2G6* pathogenic deep intronic variants are very scarce(51,53). Our literature search identified only two reported cases: c.2035-274 G>A in a PARK14-linked dystonia-parkinsonism patient(53) and c.2035-926 G>A in an INAD patient(51). Notably, all three deep intronic variants, including the SSCV identified here, are located within intron 14 of *PLA2G6*, and all contribute to the activation of a cryptic exon leading to a frameshift (Additional file 1: Figure S7).

### ● Correction of aberrant splicing induced by SSCV with ASO modality

We next investigated whether abnormal splicing identified by CrosSplice can be corrected with currently available therapeutic strategies. We selected *PLA2G6* mutation c.2034+355 G>A for experimental validation. Although PLAN is a life-threatening disease, no effective therapies are available to date, despite the preclinical research and some clinical trials testing nutritional, pharmacological and gene-based interventions(50,54). Seven ASOs (18-20 nt), fully modified with 2’MOE and PS linkages, were designed to target the SSCV and predicted ESEs (ASO sequences are shown in Table 1, and the design is illustrated in Figure 5a). To evaluate their efficacy, we conducted minigene assays in HEK293T cells by co-transfecting each ASOs with minigene constructs harbouring either wild type (WT) or mutant type (Mt) sequences. Among seven candidates, ASO_02, ASO_03 and ASO_04, which were designed to bind directly to the SSCV, consistently corrected the aberrant splicing pattern induced by the mutant allele across repeated experiments (Figure 5b,c). ASO_07 exhibited moderate corrective effects though it was designed to target ESEs distal to the SSCV. In contrast, ASO_01 showed no detectable difference compared with control samples. These findings indicate that the c.2034+355 G>A SSCV activates a cryptic exon, and its inclusion can be effectively suppressed by ASO treatment.

**Figure 5.**
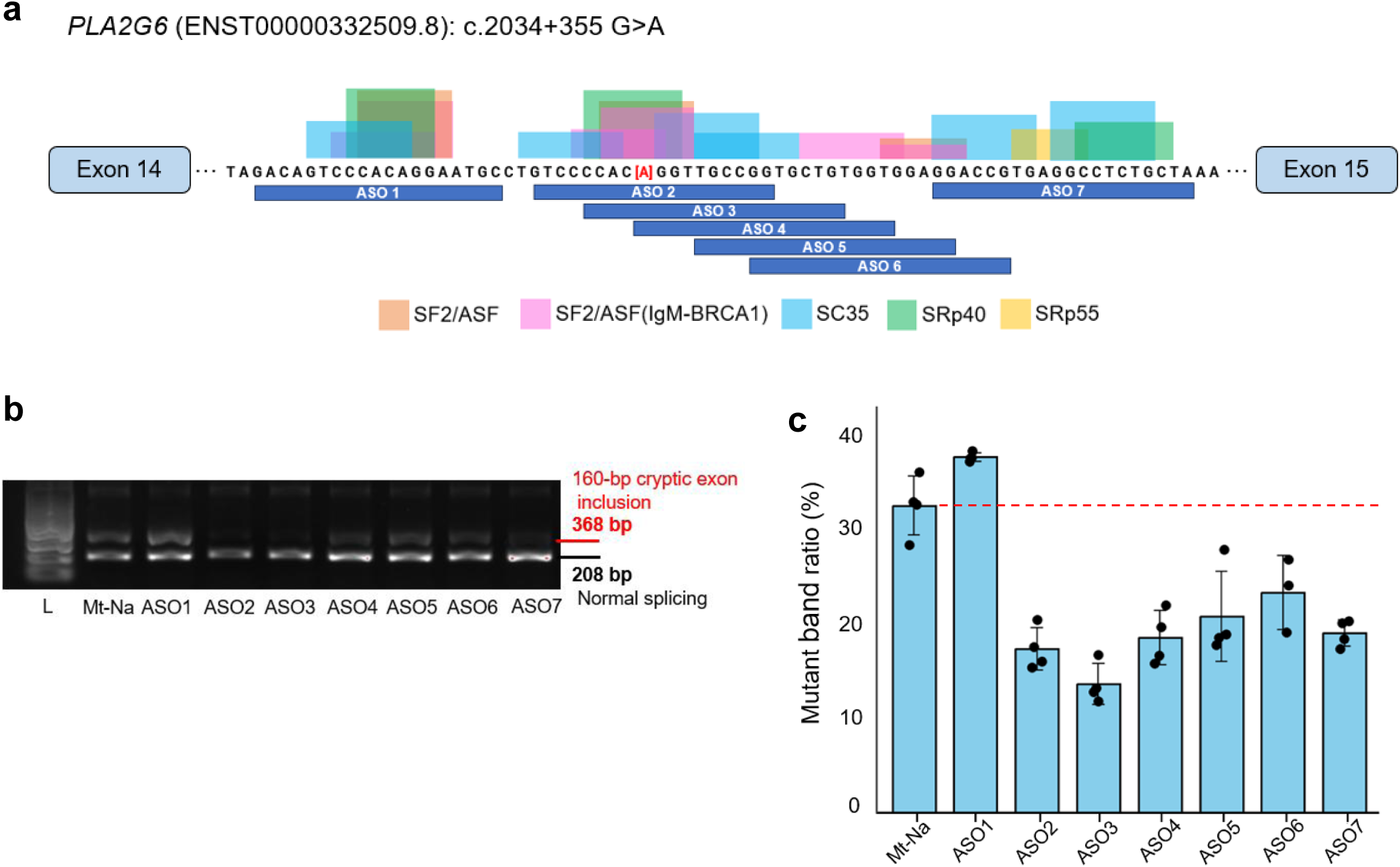
Therapeutic correction of SSCV-induced splicing alteration by ASOs. (a) Schematic of the ASO design targeting the variant and SR protein binding motifs. The motifs were identified using ESEfinder^15^ and modified for clarity. Red text enclosed in square blankets indicates the variant (Alt allele). (b)(c) Effects of seven ASOs evaluated by minigene assay with co-transfection. “Mt-Na” represents the control sample transfected with the mutant minigene construct without ASO treatment. Splicing alteration resulting in cryptic exon inclusion (368-bp band) was substantially suppressed in ASO-treated samples, particularly in ASO_2, ASO_3 and ASO_4, where the mutant band ratio was markedly reduced compared to Mt-Na. Band intensities were quantified using Image Studio 6.0 (LI-COR Biosciences). In (b), the red dashed line indicates the mean mutant band ratio of the Mt-Na group. Each bar represents the mean ± standard deviation (SD) from independent biological experiments (n=4 for each ASO, except ASO_6, which was repeated three times).

## Discussion

Several methodologies have been developed to identify SSCVs, each offering distinct strengths and limitations. sQTL-based methods provide robust population-level insights but are underpowered to detect rare splicing events. Machine learning-based predictors have substantially improved(15,17,19,55), yet their accuracy remains imperfect. Several tools have been developed to link genomic variants with RNA-based splice junctions on a per-sample basis, but their demonstrated utility has thus far been largely confined to analyses of somatic mutations in cancer(56–58). In parallel, we have recently proposed a method that identifies SSCVs solely from RNA-seq data, enabling large-scale screening from repositories such as the Sequence Read Archive(22). Though powerful for high-throughput discovery, the sensitivity of this method is not yet comprehensive.

Building upon these advances, CrosSplice achieves further improvement through its unique design, which employs machine-learning predictions to pinpoint candidate SSCVs with their associated splicing junctions, and then reinforces these predictions through statistical association testing, thereby ensuring high confidence even for rare events. Another key feature of CrosSplice is its integration of cross-tissue transcriptome data, enabling the detection of splicing events from genes expressed in only a limited number of tissues and thus providing more comprehensive coverage with improved sensitivity. As expected, the range of SSCVs detectable by CrosSplice is broader in the multi-tissue setting than in the single-tissue setting, and sensitivity improves with the number of tissues analysed. In practice, however, many available datasets are not multi-tissue but instead comprise only data from easily accessible tissues, such as blood and skin(59). Although sensitivity is inevitably reduced in the single-tissue setting, CrosSplice was still able to detect SSCVs with a reasonable degree of confidence, indicating that this approach remains applicable to such datasets for future expansion. In addition, while we adopted SpliceAI as the predictive engine for nominating primary novel SJs in this study, other emerging models, such as Pangolin or SpliceTransformer, can be incorporated in future applications.

In this study, we focused on a deep intronic variant in *PLA2G6* (c.2034+355 G>A) and demonstrated in vitro that the SSCV induced the expected splicing alteration, which was effectively corrected by splice-switching ASOs. Most *PLA2G6* mutations previously reported as the genetic cause of PLAN were located within exonic regions or at exon-intron boundaries(35,52,60–63), with a few cases (Cavestro et al.(53) and Borja et al.(51)) reporting a second pathogenic allele in deep intronic regions. This likely reflects the genome analysis methods employed in previous studies, in which whole exome sequencing (WES) was predominantly utilized. Many genetic disorders, including PLAN, still have a substantial proportion of cases remaining genetically undiagnosed, which may be attributable to overlooked pathogenic deep intronic variants(5). Moreover, deep intronic variants are considered more amenable to ASO therapeutics than others(5). In this context, the SSCV catalogue generated by CrosSplice, which encompasses a large number of potentially pathogenic variants located in deep introns, is expected to serve as a valuable resource for therapeutic targets of rare and intractable diseases.

There are still several limitations in the current CrosSplice pipeline. Although SSCVs identified by CrosSplice are generally considered to induce abnormal splicing, they do not necessarily possess clinical pathogenicity. This discrepancy may arise for several reasons. For example, the penetrance of splicing alterations caused by SSCVs may be incomplete, and the residual functional protein may be sufficient to sustain the essential biological functions. Moreover, SSCVs do not always result in simple loss-of-function transcripts. In some cases, pseudoexon inclusion may introduce novel domains, potentially giving rise to gain-of-function proteins(22). At present, each variant interpretation still requires careful literature review. However, the integration of artificial intelligence is expected to play an increasingly important role in this process(64).

Beyond interpretation, experimental validation using minigene or cell-based assays and the design of ASO also remain largely manual, making them labor-intensive and potentially inconsistent across studies. Recently, artificial intelligence approaches have enabled partial automation of these processes, exemplified by tools such as ASOptimizer(65). Nevertheless, their current performance is still limited by restricted training data, poor interpretability, and uncertainties in predicting clinical outcomes. Looking ahead, extending automation beyond annotation and interpretation to include ASO design and even experimental validation steps could pave the way for a fully integrated and efficient workflow.

## Conclusion

CrosSplice substantially extends the detectable landscape of SSCVs by capturing rare and tissue-specific events that are beyond the reach of conventional approaches. The resulting SSCV catalogue, enriched for deep intronic variants that are highly amenable to ASO-mediated splice modulation, provides a valuable resource for therapeutic target discovery in rare and intractable diseases and facilitates the development of ASO therapies for those conditions.

## Abbreviations

AF: Allele frequency
ASO: Antisense oligonucleotide
ESE: Exonic splicing enhancer
FDR: False discovery rate
GTEx: Genotype-Tissue-Expression
INAD: Infantile neuroaxonal dystrophy
2’ MOE: 2’-O-Methoxyethyl
Mt: Mutant type
NMD: Nonsense-mediated decay
PLAN: PLA2G6-associated neurodegeneration disorders
PS: Phosphorothioate
PTC: Premature termination codon
SCA: Spinocerebellar ataxia
SNV: Single nucleotide variants
SJ: Splicing junction
sQTL: Splicing quantitative trait loci
SS: Splice-site
SSCV: Splice-site creating variant
WES: Whole exome sequencing
WGS: Whole genome sequencing
WT: Wild type

## Declaration

### ● Ethics approval and consent to participate

Not applicable.

### ● Consent for publication

Not applicable.

### ● Availability of data and materials

The GTEx transcriptome sequencing data and whole-genome genotype calls in VCF format were obtained from the Sequence Read Archive. All codes used in this study are available at the CrosSplice GitHub repository (https://github.com/yuki-yano10/CrosSplice/). The list of identified SSCVs is provided in Additional file 2 (.xlsx).

### ● Authors’ contributions

YS designed the study. YY and AO developed the software of CrosSplice. YY performed the pipeline on the GTEx datasets with assistance from AO, and analysed and interpreted the list of SSCVs. YY, MO and TN conducted all the vitro experiments, including minigene assays and design and administration of ASO. YY wrote the manuscript with assistance from TN, RNM and YS.

## ● Acknowledgements

We thank Marlen Lauffer and her colleagues at DCRT for allowing us to use their gene list. The supercomputing resources were provided by the Human Genome Center, the Institute of Medical Science, The University of Tokyo. We would like to thank the GTEx Consortium for providing the data on which this study is based.

## Supplementary information

### Supplementary figures

**Figure S1.**
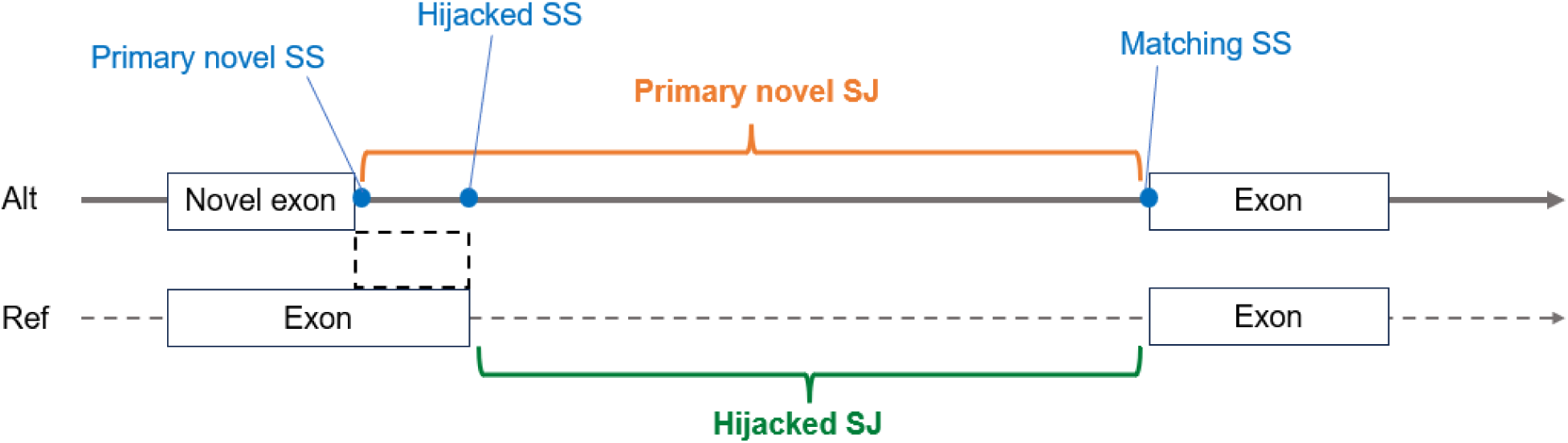
Schematic illustration of the definition of hijacked splice site (SS), matching SS, hijacked splice junction (SJ) and primary novel SJ. The example shows a donor creating variant on the positive strand that results in partial exon loss. Primary novel SS is defined as a splice site newly created by an SSCV, while matching SS denotes its partner splice-site (the nearest upstream donor site in the case of an acceptor gain, or the nearest downstream acceptor site in the case of a donor gain). Hijacked SS is the original splice-site that had been paired with the matching SS but displaced by the primary novel SS. Primary novel SJ is defined as the region between the primary novel SS and matching SS, whereas hijacked SJ is the region between the hijacked SS and the matching SS.

**Figure S2.**
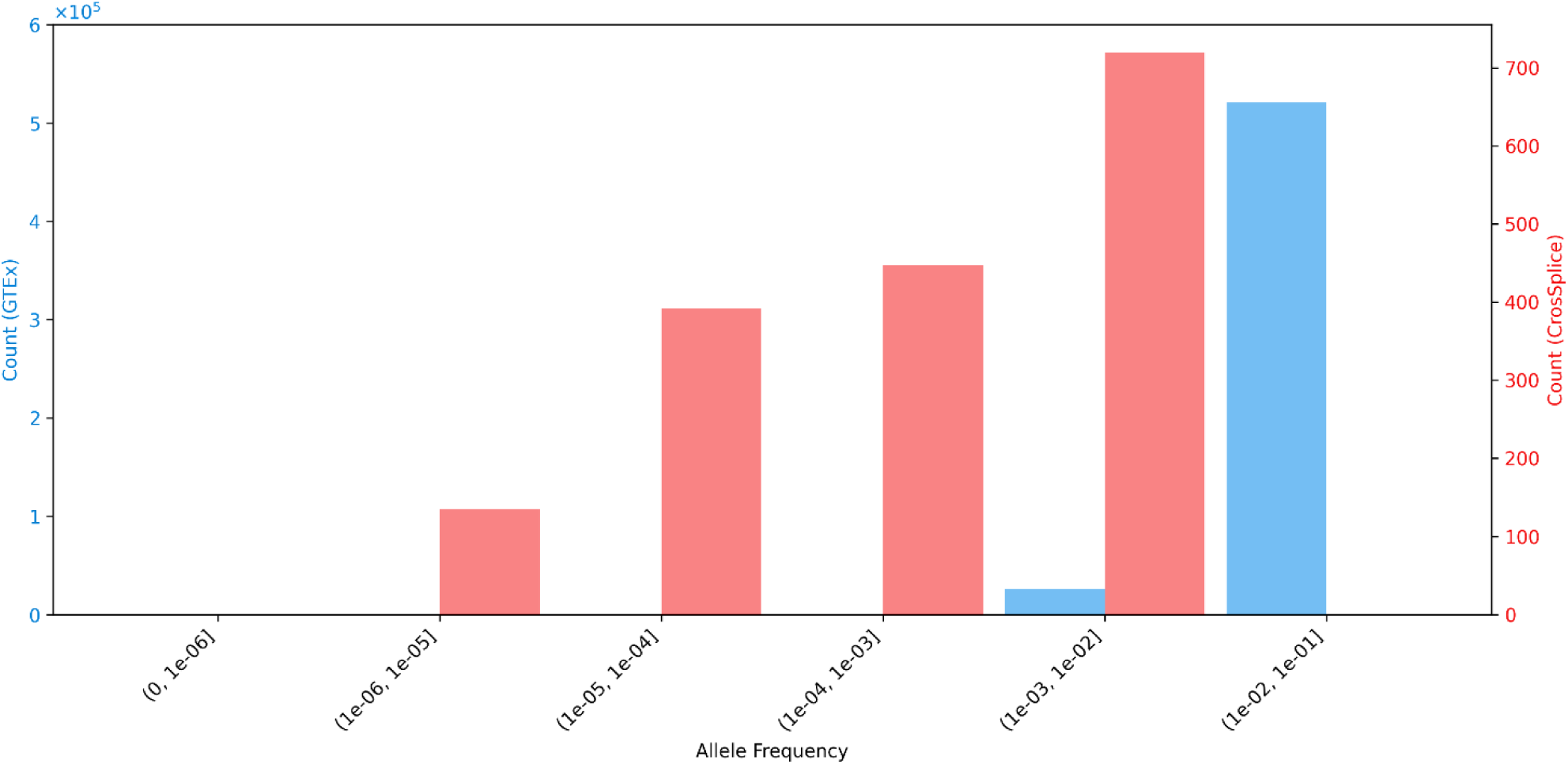
Distribution of allele frequencies of GTEx v8 sQTL SNVs and CrosSplice-identified SSCVs. Distribution of minor allele frequencies of GTEx v8 sQTL SNVs and gnomADg allele frequencies of CrosSplice-identified SSCVs are compared. Since CrosSplice analysed only SNVs with allele frequencies ≤ 0.01, this figure shows the allele frequency distributions of SSCVs (CrosSplice) and sQTL SNVs (GTEx) within this range.

**Figure S3.**
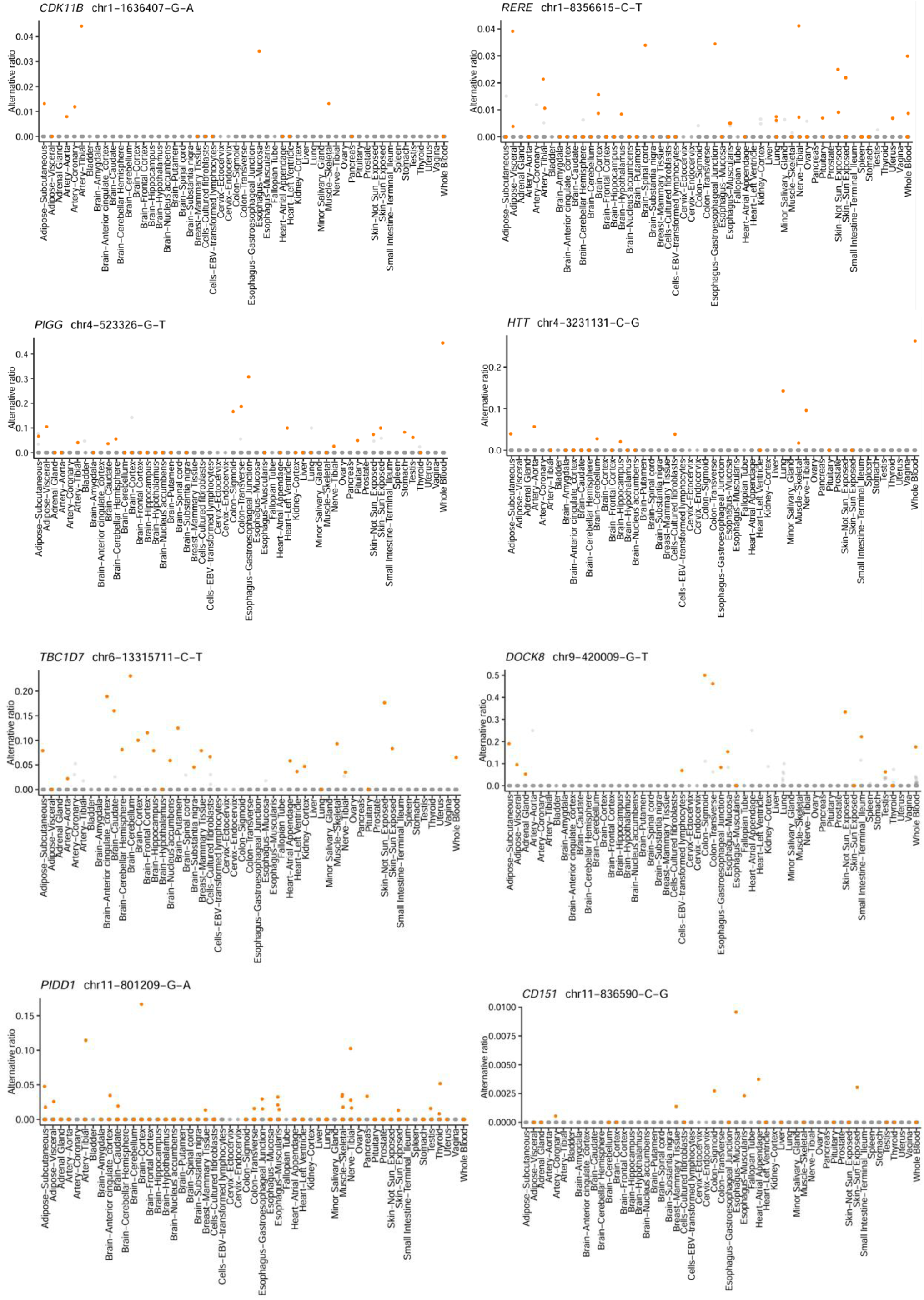
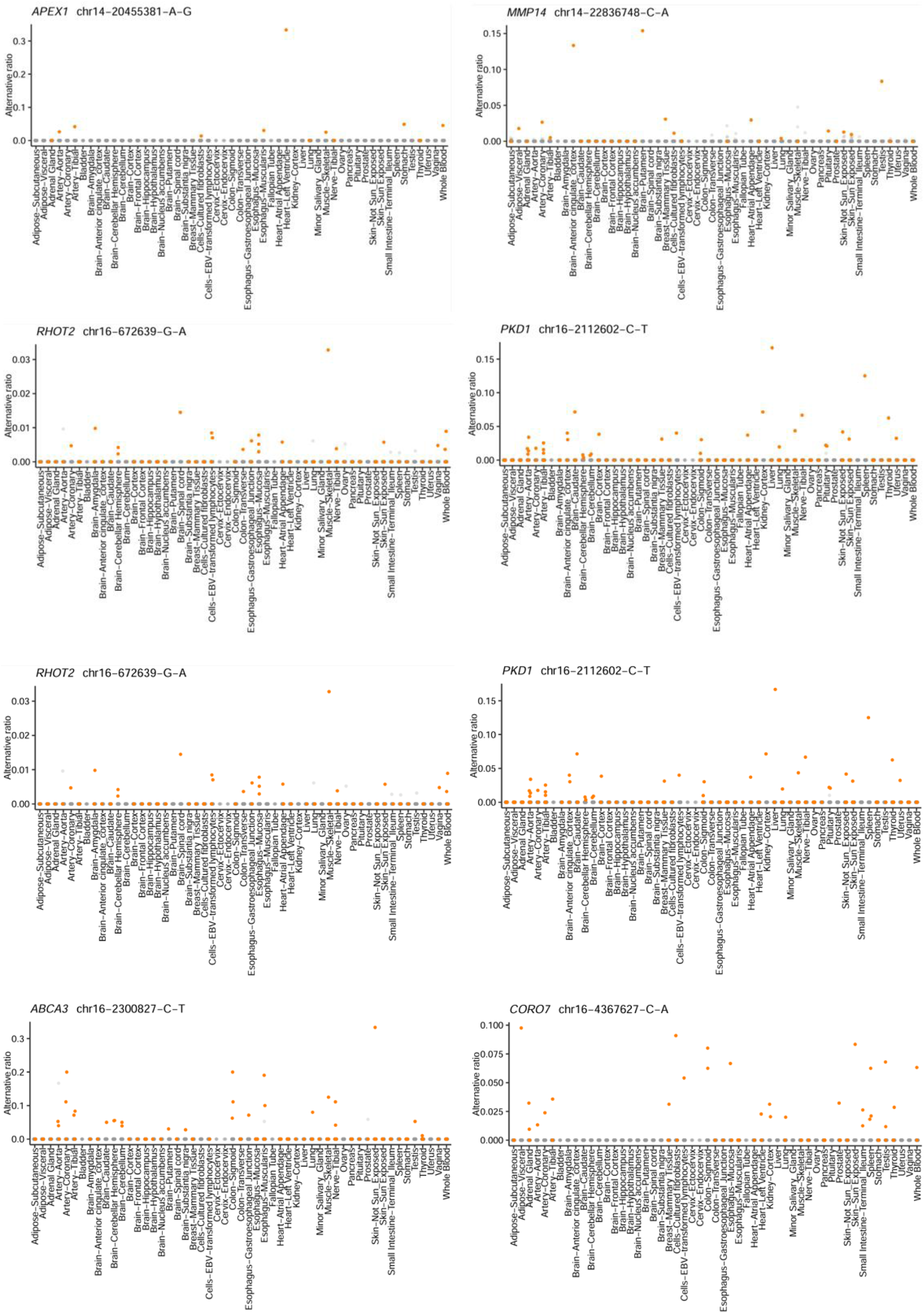
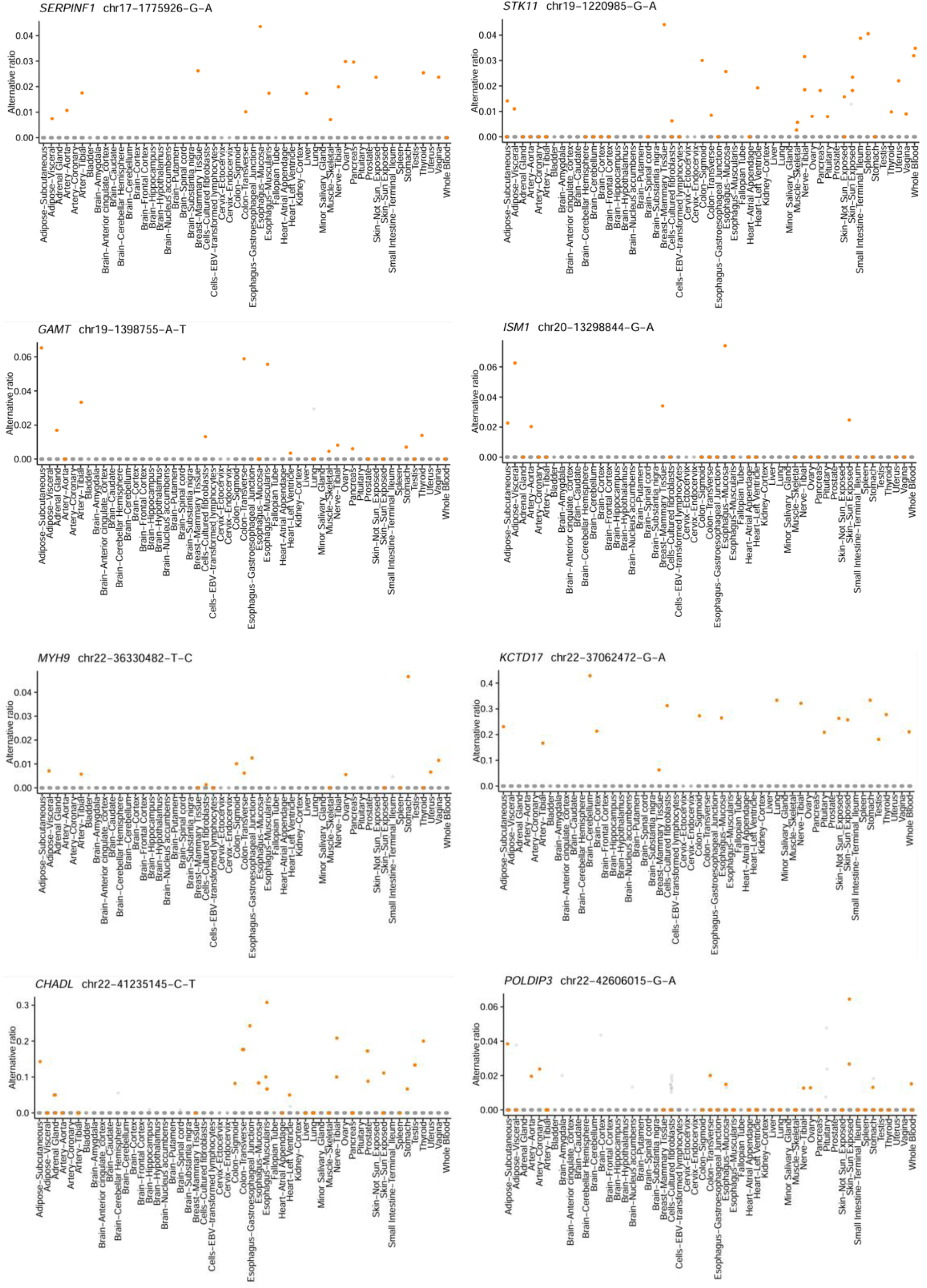
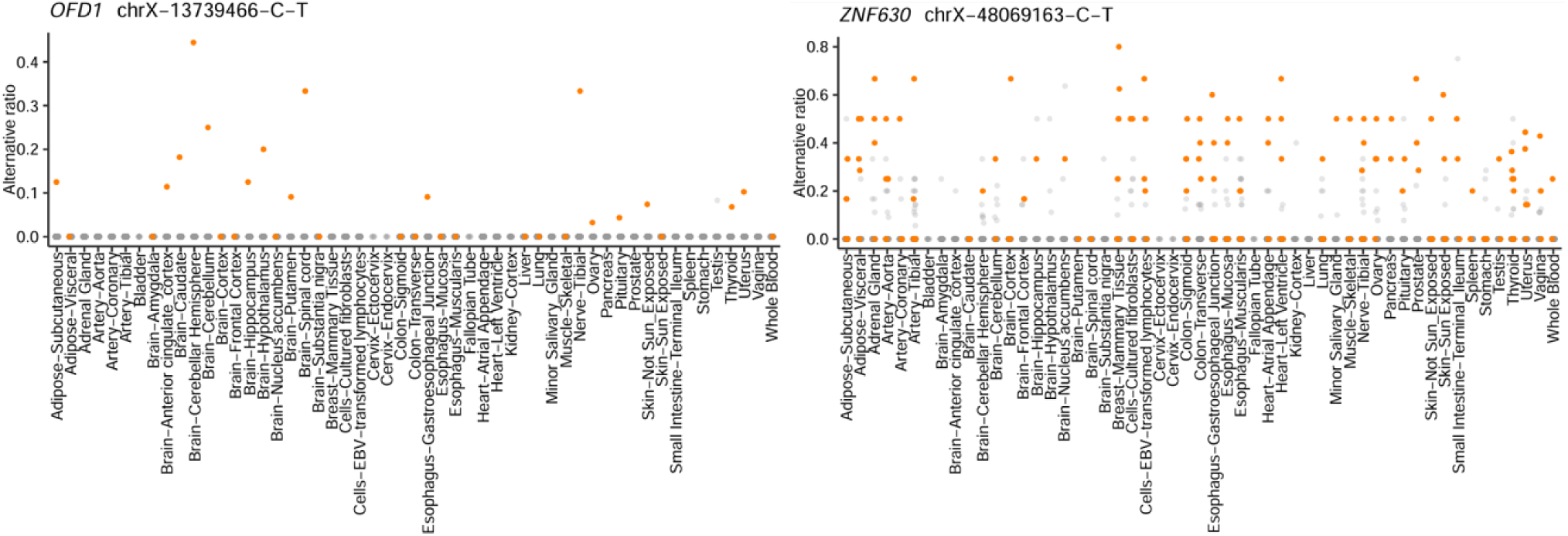
26 SSCVs with low SpliceAI delta scores (< 0.2 range) exhibiting high CrosSplice scores (> 300). Alternative ratios across tissues for the 26 SSCVs with low SpliceAI delta score and high CrosSplice score. Orange dots indicate alternative ratios in samples harbouring the variant, and grey dots represent those in samples without the variant.

**Figure S4.**
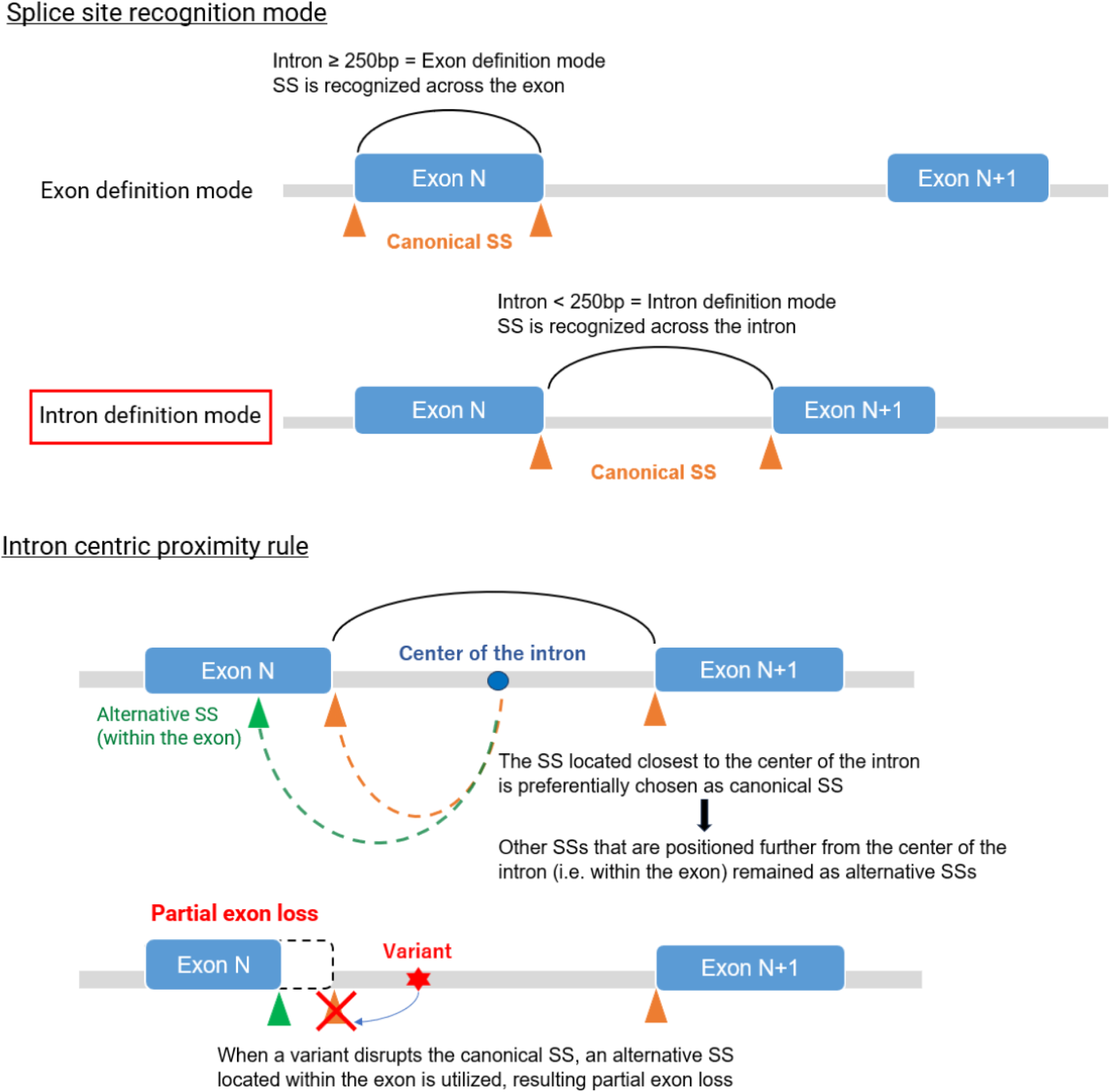
Schematic diagrams illustrating splice site recognition modes and the intron centric proximity rule. Above: Splice sites are recognized through two distinct modes depending on the intron length: exon definition and intron definition. When the intron is longer than 250bp, the exon definition mode is adopted, whereas with shorter introns splice sites are recognized via the intron definition mode. Intron definition mode emphasizes the intron centric proximity rule (below). Below: Under this rule, the splice site located closest to the center of the intron is preferentially selected among multiple candidates, leaving alternative splice sites within the exon. When a variant weakens a canonical splice site, an alternative splice site within the exon is often utilized, resulting in partial exon loss.

**Figure S5.**
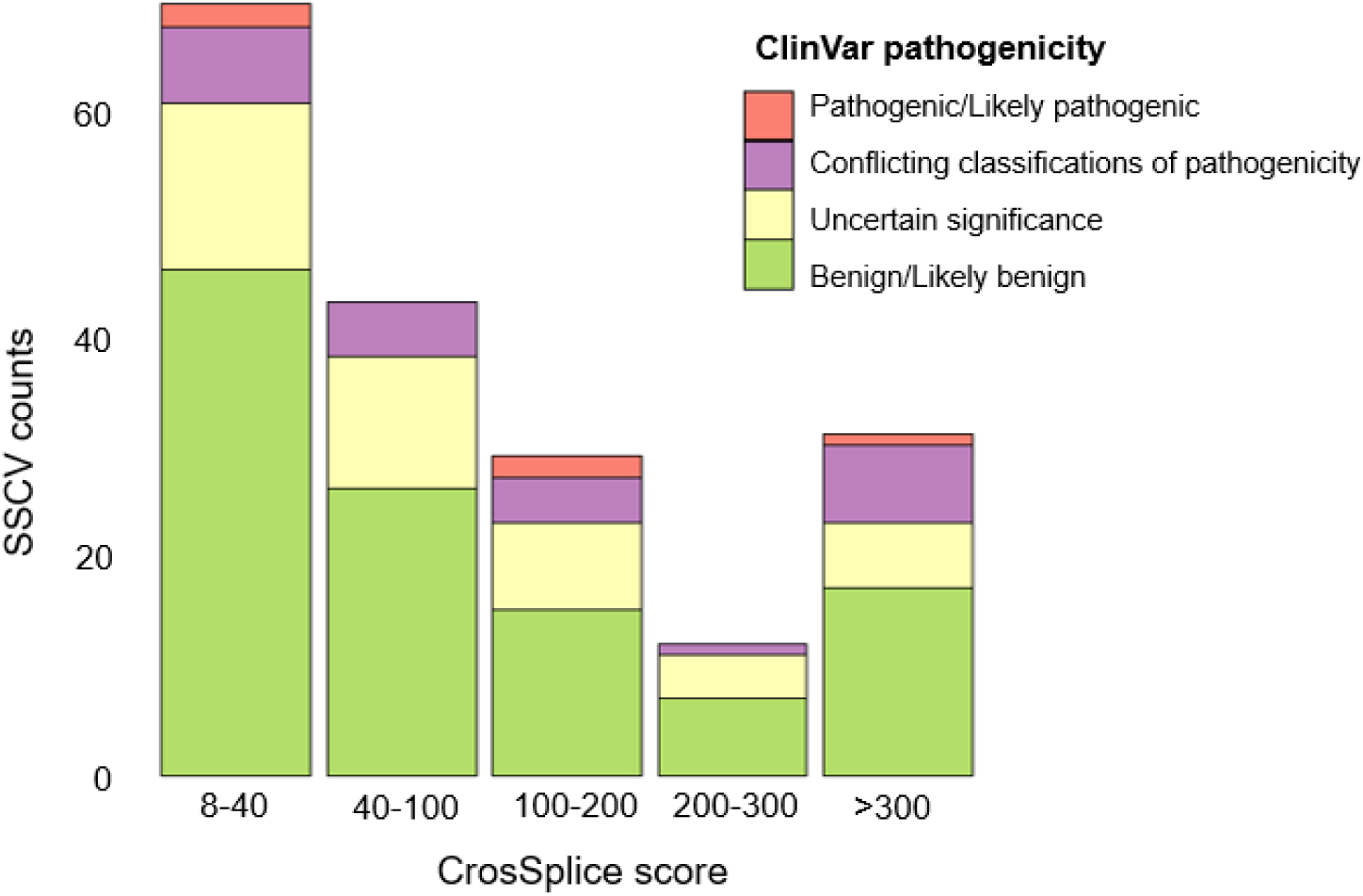
Distribution of ClinVar pathogenicity classifications across CrosSplice score ranges for identified SSCVs. The number of SSCVs within each CrosSplice score range is shown on the y-axis. ClinVar pathogenicity classifications are indicated by the fill colors.

**Figure S6.**
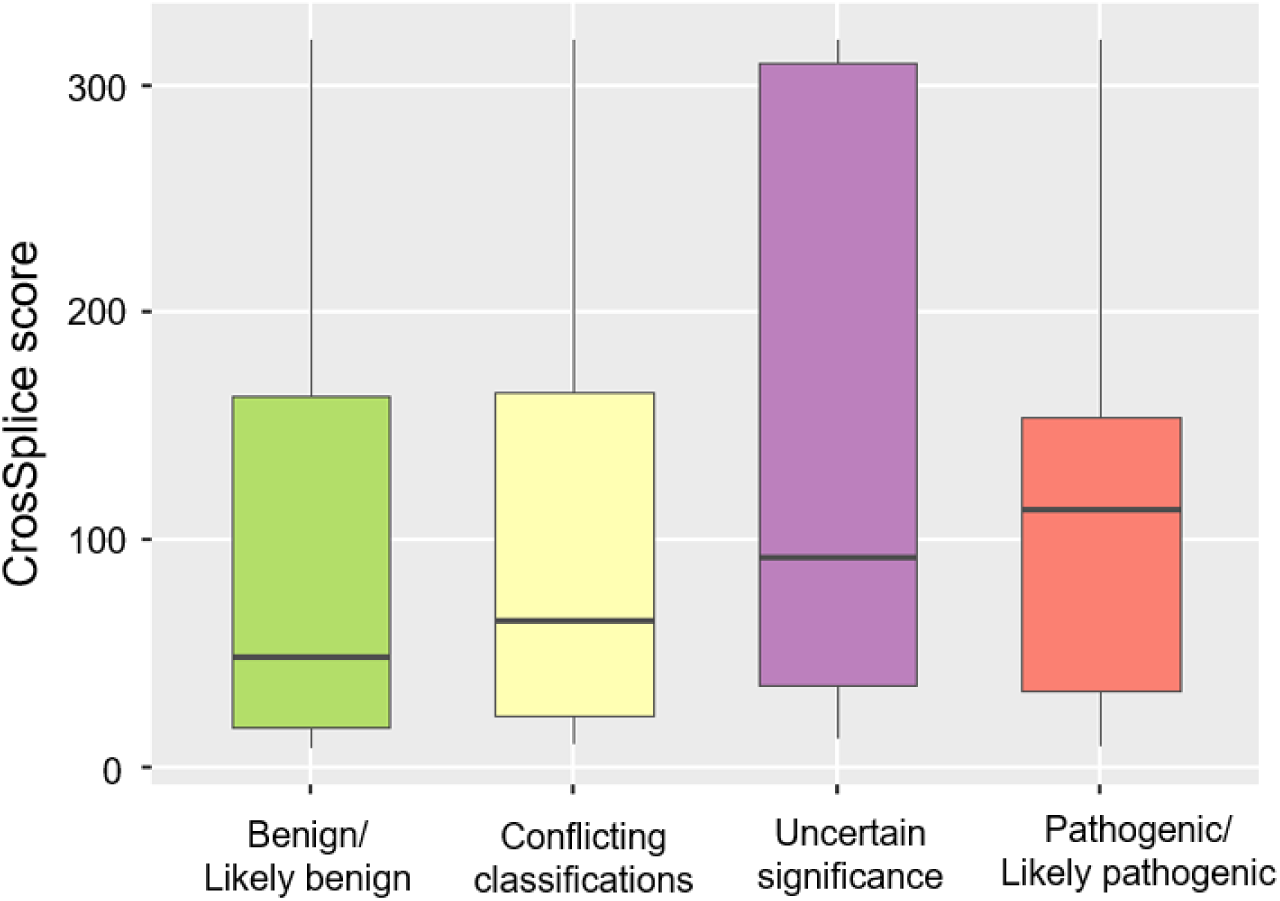
Boxplot showing the distribution of CrosSplice scores across ClinVar pathogenicity classifications. CrosSplice scores tended to be higher in pathogenic/likely pathogenic variants and lower in benign/likely benign variants; however no statistically significant differences were observed among the pathogenicity groups (Kruskal-Wallis test and pairwise Wilcoxon test, ggplot2 package in R).

**Figure S7.**
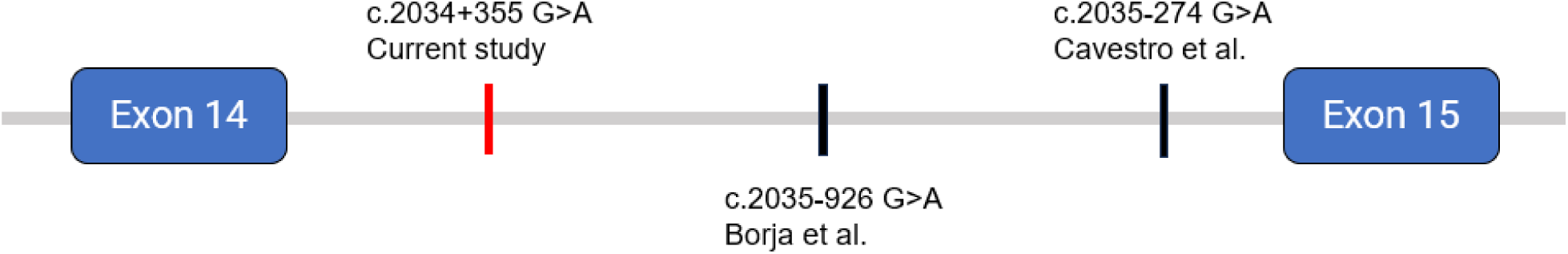
Schematic representation of three deep intronic variants in *PLA2G6* gene. The SSCV identified in the current study (c.2034+355 G>A) and two previously reported variants (c.2035–926 G>A by Borja et al.39 and c.2035–274 G>A by Cavestro et al.41) are all located within intron 14.

### Supplementary tables

**Table S1. 1,743 significant SSCVs identified by CrosSplice.**

Submitted in Excel format.

This table summarizes all significant SSCVs detected by CrosSplice in the GTEx v7 dataset. Each row represents an SSCV meeting the CrosSplice significance threshold (score ≥ 8). The table includes genomic coordinates, gene symbols, HGVS c. notations, SpliceAI delta scores, gnomAD allele frequencies, ClinVar annotations, splicing junction information, and predicted splicing consequences. Key, a genomic coordinate with a gene symbol (formatted as “chr_pos_Ref_Alt_Gene”); SpliceAI DS, SpliceAI delta score (maximum of DS_AG and DS_DG); gnomADg AF, allele frequency in gnomAD v3.1.2; SSCV type, donor or acceptor gain; Primary novel SJ, newly created splice junction generated by SSCV; Hijacked SJ, canonical splice junction displaced by SSCV; Phenotype, known disease association from the DCRT list, primarily based on OMIM/ORPHA. HGVS c. notations are provided primarily based on MANE Select transcripts (GENCODE v39).

**Table S2.**
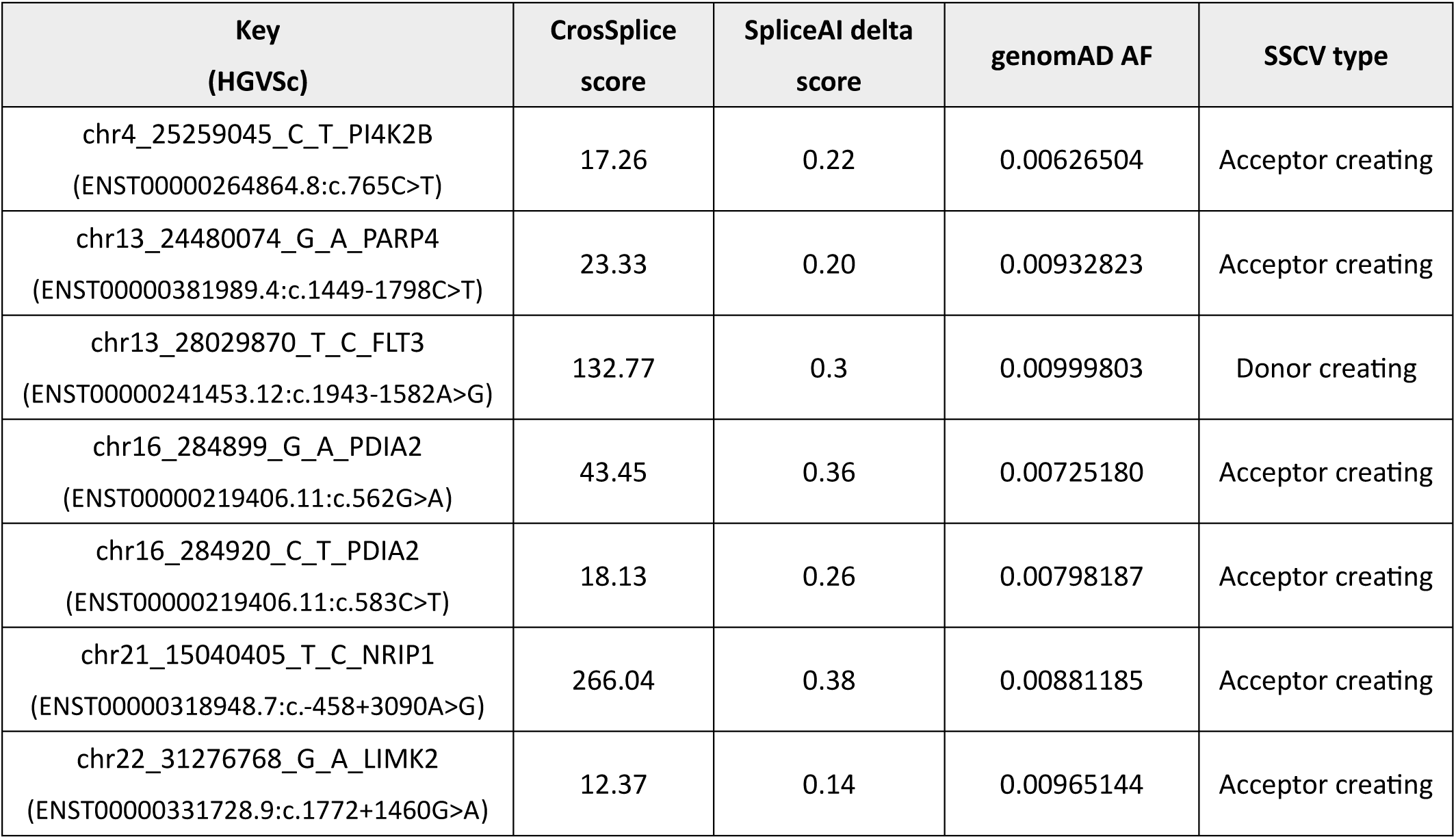
Seven overlapping SSCVs between GTEx v8 sQTL SNVs and CrosSplice-identified SSCVs. Comparison between 1,875,911 SNVs identified as sQTLs by LeafCutter in the GTEx v8 dataset and 1,743 SSCVs identified by CrosSplice revealed only seven overlapping SSCVs. Key represents a genomic coordinate with a gene symbol (formatted as “chr_pos_Ref_Alt_Gene”). HGVS c. notations (below) are provided primarily based on MANE Select transcripts (GENCODE v39). SpliceAI DS, SpliceAI delta score (maximum of DS_AG and DS_DG); gnomADg AF, allele frequency in gnomAD v3.1.2; SSCV type, donor or acceptor gain.

| Key<br>(HGVS) | CrosSplice<br>score | SpliceAI delta<br>score | gnomAD AF | SSCV type |
| --- | --- | --- | --- | --- |
| chr4_25259045_C_T_PI4K2B<br>(ENST00000264864.8:c.765C>T) | 17.26 | 0.22 | 0.00626504 | Acceptor creating |
| chr13_24480074_G_A_PARP4<br>(ENST00000381989.4:c.1449-1798C>T) | 23.33 | 0.20 | 0.00932823 | Acceptor creating |
| chr13_28029870_T_C_FLT3<br>(ENST00000241453.12:c.1943-1582A>G) | 132.77 | 0.3 | 0.00999803 | Donor creating |
| chr16_284899_G_A_PDIA2<br>(ENST00000219406.11:c.562G>A) | 43.45 | 0.36 | 0.00725180 | Acceptor creating |
| chr16_284920_C_T_PDIA2<br>(ENST00000219406.11:c.583C>T) | 18.13 | 0.26 | 0.00798187 | Acceptor creating |
| chr21_15040405_T_C_NRIP1<br>(ENST00000318948.7:c.-458+3090A>G) | 266.04 | 0.38 | 0.00881185 | Acceptor creating |
| chr22_31276768_G_A_LIMK2<br>(ENST00000331728.9:c.1772+1460G>A) | 12.37 | 0.14 | 0.00965144 | Acceptor creating |

**Table S3.** False discovery rate with various thresholds. False discovery rates (FDRs) were estimated by permutation tests at various CrosSplice score thresholds.

| Threshold (CrosSplice score) | FDR (%) |
| --- | --- |
| 1 | 18.8 |
| 2 | 12.1 |
| 4 | 8.1 |
| 6 | 6.3 |
| 8 | 5.2 |
| 10 | 4.3 |
| 12 | 3.6 |
| 14 | 3.2 |
| 16 | 2.8 |
| 18 | 2.5 |
| 20 | 2.2 |
| 30 | 1.3 |
| 40 | 0.9 |

